# Dietary nitrate intake and net nitrite-generating capacity of the oral microbiome interact to enhance cardiometabolic health: Results from the Oral Infections Glucose Intolerance and Insulin Resistance Study (ORIGINS)

**DOI:** 10.1101/2024.04.10.24305636

**Authors:** Charlene E. Goh, Bruno Bohn, Jeanine M. Genkinger, Rebecca Molinsky, Sumith Roy, Bruce J. Paster, Ching-Yuan Chen, Melana Yuzefpolskaya, Paolo C. Colombo, Michael Rosenbaum, Rob Knight, Moïse Desvarieux, Panos N. Papapanou, David R. Jacobs, Ryan T. Demmer

**Author notes:** **Corresponding authors: Charlene Goh, BDS, DrPH** 9 Lower Kent Ridge Road, Singapore 119085, Singapore **Ryan Demmer, PhD** 205 3^rd^ Avenue SW Rochester, MN 55905.

## Abstract

**Background:** We investigated the association between dietary nitrate intake and early clinical cardiometabolic risk biomarkers, and explored whether the oral microbiome modifies the association between dietary nitrate intake and cardiometabolic biomarkers.

**Methods:** Cross-sectional data from 668 (mean [SD] age 31 [9] years, 73% women) participants was analyzed. Dietary nitrate intakes and alternative healthy eating index (AHEI) scores were calculated from food frequency questionnaire responses and a validated US food database. Subgingival 16S rRNA microbial genes (Illumina, MiSeq) were sequenced, and PICRUSt2 estimated metagenomic content. The Microbiome Induced Nitric oxide Enrichment Score (MINES) was calculated as a microbial gene abundance ratio representing enhanced net capacity for NO generation. Cardiometabolic risk biomarkers included systolic and diastolic blood pressure, HbA1c, glucose, insulin, and insulin resistance (HOMA-IR), and were regressed on nitrate intake tertiles in adjusted multivariable linear models.

**Results:** Mean nitrate intake was 190[171] mg/day. Higher nitrate intake was associated with lower insulin, and HOMA-IR but particularly among participants with low abundance of oral nitrite enriching bacteria. For example, among participants with a low MINES, mean insulin[95%CI] levels in high vs. low dietary nitrate consumers were 5.8[5.3,6.5] vs. 6.8[6.2,7.5] (p=0.004) while respective insulin levels were 6.0[5.4,6.6] vs. 5.9[5.3,6.5] (p=0.76) among partcipants with high MINES (interaction p=0.02).

**Conclusion:** Higher dietary nitrate intake was only associated with lower insulin and insulin resistance among individuals with reduced capacity for oral microbe-induced nitrite enrichment. These findings have implications for future precision medicine-oriented approaches that might consider assessing the oral microbiome prior to enrollment into dietary interventions or making dietary recommendations.

**Clinical Perspective:** *What is new?:*

- In this population-based study we identified an interaction between dietary nitrate intake and oral nitrite enriching bacteria on cardiometabolic outcomes.
- Higher dietary nitrate intake was associated with lower insulin and insulin resistance *only* among participants with low abundance of oral nitrite enriching bacteria.
- This study suggests that cardiometabolic benefits of nitrate consumption might depend on the host microbiome’s capacity to metabolize nitrates.

*What are the clinical implications?:*

- Among people with low microbiome capacity for nitrate metabolism, higher levels of nitrate might be necessary to realize cardiometabolic benefits.
- Lack of microbiome assessments in prior studies could partially explain inconsistent findings from previous nitrate supplementation trials and observational studies.
- Future precision-medicine oriented trials studying the effects of dietary nitrate recommendations on cardiometabolic health, should consider assessing the oral microbiome.

## INTRODUCTION

Dietary nitrate consumption (derived mainly from vegetables such as dark leafy greens and beetroot) is associated with improved cardiometabolic health in several, but not all prior studies^1–5^. However, the mechanisms underlying these potential cardiometabolic benefits are not fully understood and the source of heterogeneity of reported effects is unknown, limiting causal inference and translation into clinical and public health practice.

Prior heterogeneity in research findings could be explained by the fact that cardiometabolic benefits of nitrate consumption are partially reliant on the oral microbiome-dependent enterosalivary nitrate-nitrite-nitric oxide (NO_3_- NO_2_- NO) pathway, by which oral microbes convert dietary nitrate to nitrite (Figure 1A). NO_2_, once swallowed is made bioavailable for NO production. In turn, increased NO bioavailability improves blood flow, insulin-mediated glucose uptake^6,7^, blood pressure, and other cardiometabolic biomarkers^8^. Thus, the oral microbiome could jointly explain previous variability in study results while also providing mechanistic clues about the benefits of nitrate consumption.

**Figure 1.**
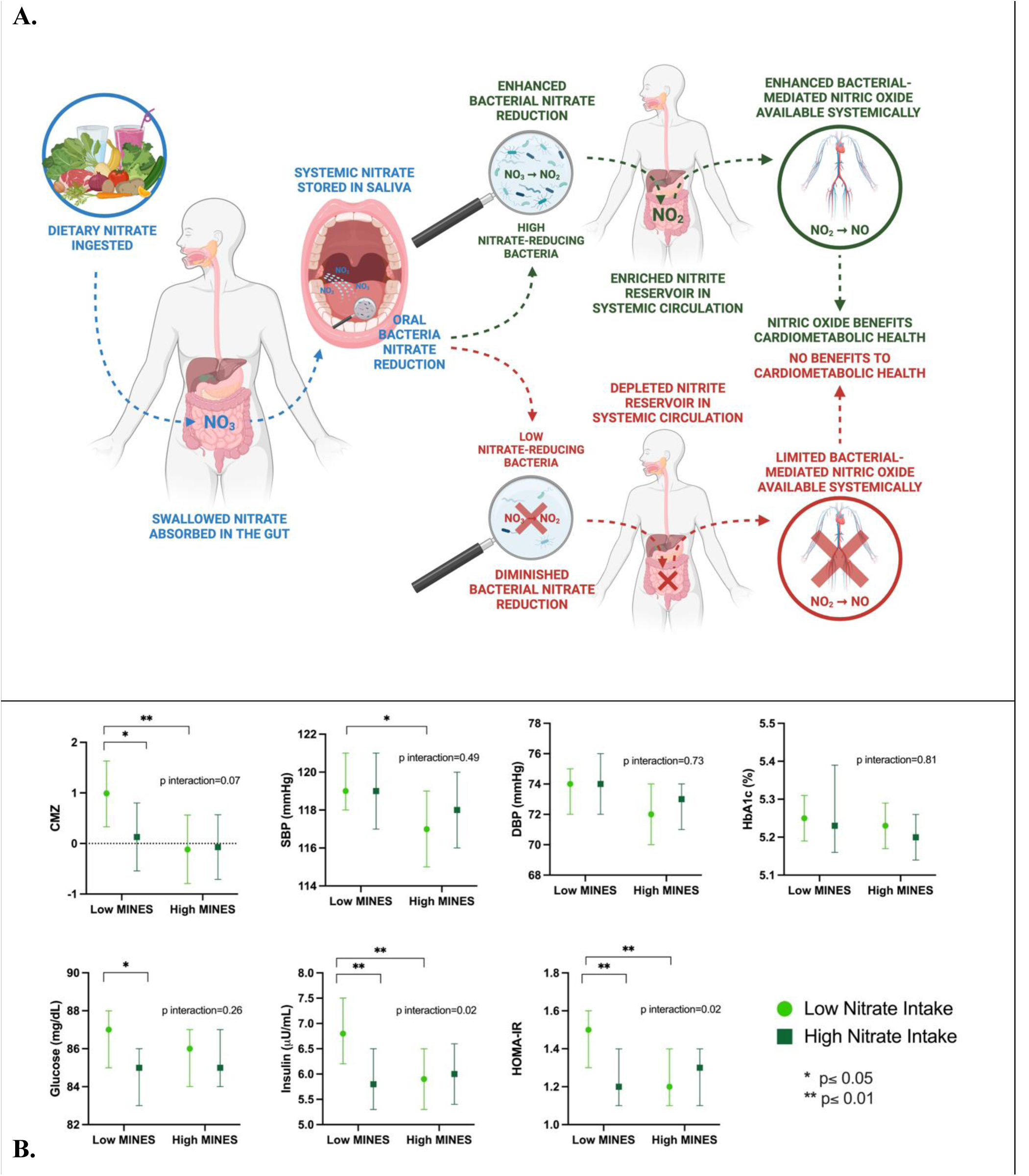
A) Conceptual overview of the process by which the oral microbiome facilitates the generation of bioavailable nitric oxide. Nitrates consumed via the diet (e.g., green leafy vegetables), or endogenously generated, are concentrated in saliva. Oral bacteria reduce salivary nitrate to nitrite in the mouth and nitrite is in turn swallowed and converted to systemically available NO via enzymatic and non-enzymatic downstream pathways. *A necessary step in this chain of events is bacterial-dependent conversion of nitrate to nitrite. This system is viewed as a “backup system” ensuring a reservoir of NO is available when NO generation from other pathways is compromised (e.g., dysfunction in endothelium-dependent vasodilation^8^). B) Mean cardiometabolic biomarker levels by Low vs. High categories of dietary nitrate intake (median split) and above vs. below median MINES. (The corresponding interaction analyses results used to generate this figure are available in Supplementary Table S4) MINES: Microbiome Induced Nitric oxide Enrichment Score was calculated as a ratio of microbial gene abundances necessary for NO vs. NH_3_ production. This score is a surrogate for enhanced net capacity for nitrite generation. These data support the concept that among individuals with low dietary nitrate intake an oral microbiome enriched for nitrite generation can ensure an adequate reservoir of NO-generating capacity and promote cardiometabolic health (i.e., the microbiome buffers against a low nitrate diet). Or, conversely, among individuals with an oral microbiome that has limited nitrite generating capacity (e.g., oral dysbiosis) a diet high in nitrates might ensure an adequate reservoir of NO-generating capacity and promote cardiometabolic health (i.e., the diet buffers against a dysbiotic microbiome).

The role of the oral microbiome in the NO_3_- NO_2_- NO pathway has been demonstrated by several experimental trials (see^1,2^ for reviews). Accordingly, small experimental studies that reduce oral nitrate reduction potential via antibacterial mouthwash report blunted blood pressure lowering effects of nitrate supplementation^9,10^. However, although both higher dietary nitrate intake and higher levels of oral nitrate-reducing capacity have been separately associated with lower cardiometabolic risk outcomes in population-based studies^1,2,11,12^, no prior population-based study has directly examined the interaction between dietary nitrate intake and the nitrate reducing/nitrite-generating capacity of the oral microbiome in relation to cardiometabolic outcomes.

The need for more epidemiological research on cardiometabolic benefits of nitrate consumption as well as synergy between the microbiome and dietary patterns was emphasized by the *2016 National Heart, Lung, and Blood Institute Workshop on Dietary Nitrate*^13^ and the *2020-2030 Strategic Plan for NIH Nutrition Research*^14^. Such studies can inform the real-world effectiveness of habitual dietary nitrate intake, which occurs at doses lower than those used in most experimental trials testing the cardiometabolic benefits of nitrate supplementation. Furthermore, understanding the association of dietary nitrate intake with early clinical biomarkers, such as elevated insulin resistance and blood pressure, prior to the onset of disease and associated pathophysiological alterations, is warranted to understand the window of opportunity for prevention and intervention. These studies can also inform whether oral microbiome assessments, and/or adjunctive therapies targeting microbial net nitrite-generating capacity, should be incorporated in future dietary nitrate supplementation studies.

The aim of this study is to investigate the association between dietary nitrate intake and early clinical cardiometabolic risk biomarkers, and to explore whether the oral microbiome modifies the association between dietary nitrate intake and cardiometabolic biomarkers. We utilize cross-sectional data from a population-based sample of predominantly young, healthy adults in the US, and hypothesize that the association between dietary nitrate intake and cardiometabolic outcomes will be modified by the abundance of net nitrite-generating bacterial genes present.

## METHODS

### Description of ORIGINS

ORIGINS is a prospective cohort study investigating the relationship between the subgingival microbial community composition and impaired glucose metabolism^15,16^. The cross-sectional data used in this study are from a subset of participants with standardized oral microbiota and dietary data collected. Inclusion criteria were: i) aged 20-55 years; ii) no diagnosis of diabetes mellitus based on participant self-report, HbA1c values <6.5% and fasting plasma glucose<126 mg/dl; iii) no self-reported history of myocardial infarction, congestive heart failure, stroke, or chronic inflammatory conditions. All participants reported not taking antibiotics in the past 30 days. The Columbia University and University of Minnesota Institutional Review Boards approved the protocol. All participants provided informed consent. Participants who completed a food frequency questionnaire (FFQ), did not have implausible food energy levels (females with food energy values ≤ 600 or ≥ 6,000 and males with food energy values ≤ 800 or ≥ 8,000 were excluded as commonly done by other studies^17^), had available 16S rRNA microbial data from subgingival plaque samples, and were not missing important baseline risk factors or outcome data were included in this analysis, producing a study population of n=668 participants (out of 800 participants of ORIGINS Wave 2) for these analyses.

### Operationalization of Dietary Nitrate Intake Exposure

Baseline food and nutrient consumption were assessed via the National Cancer Institute Diet History Questionnaire 1 (DHQ-1) which queries frequency of consumption and portion size for 124 food items^18^. Previous studies demonstrate the validity and reproducibility of the DHQ in a multi-ethnic population similar to the ORIGINS population^19^.

Dietary nitrate intake was determined by summing the nitrate content of each food item. Nitrate values were obtained from a US nitrate food composition database developed and validated utilizing an earlier version of the DHQ-1^20^. Nitrate content of foods items not available in the database were obtained from other best available reference databases or sources^21^, derived from closely related foods or the main ingredient of that item, or calculated using standardized recipes. Further details regarding the calculation methodology, reference databases and nitrate values used are provided in the Supplemental Materials and Table S1.

The National Cancer Institute’s Diet*Calc software [Diet*Calc Analysis Program, Version 1.5.0. National Cancer Institute] was used together with the nitrate values for each food item to calculate the total daily nitrate intake for each individual based on the FFQ responses, and adjusted for total energy intake (kilocalories) using the residual method^22^, to create an energy-adjusted nitrate intake variable.

### Cardiometabolic risk biomarkers

Plasma glucose, hemoglobin A1c (HbA1c), and insulin were measured using standard methods following an overnight fast ^15,23^. The Homeostasis Model Assessment for Insulin Resistance (HOMA-IR) was used to determine insulin resistance, calculated from fasting insulin and glucose levels^24^. Seated resting systolic and diastolic blood pressures were measured in triplicate, and the last two measurements were averaged.

Systolic and diastolic blood pressure, HbA1c, fasting plasma glucose, insulin, and HOMA-IR Z-scores were calculated for each cardiometabolic risk variable by subtracting the study population mean value from the individual participant value, divided by the population standard deviation. Insulin resistance and plasma insulin were natural log-transformed to address their skewed distributions before standardization into Z-scores. Overall composite cardiometabolic risk was calculated as the average of the above individual cardiometabolic risk variable Z-scores, with a higher composite cardiometabolic Z-score (CMZ) indicating worse cardiometabolic health.

### Demographic, Anthropometric and Behavioral Risk Factors

Trained research assistants collected data on additional cardiometabolic risk factors as previously described^15^. Questionnaires assessed information on age, sex, race/ethnicity (non-Hispanic Black, non-Hispanic White, Hispanic, Other), educational level (<Bachelor’s degree, Bachelor’s degree, >Bachelor’s degree), and smoking status (current, former, or never smoking). Body mass index (BMI) was calculated as weight (in kilograms)/height (meters^2^) from in-person physical assessments. Periodontal status was evaluated through clinical oral examinations and classified according to the Centers for Disease Control and Prevention/American Academy of Periodontology (CDC/AAP) diagnosis classification (None/Mild, Moderate/Severe)^25^.

The Alternative Healthy Eating Index-2010 (AHEI), a dietary score created based on foods and nutrients predictive of chronic disease risk^26^, was calculated from food frequency data as previously described^16^. The index consists of eleven dietary component groups (vegetables, fruits, whole grains, nuts and vegetable protein, red/processed meat, sugar-sweetened beverages and fruit juice, trans fats, polyunsaturated fats, long-chain fatty acids, sodium, and alcohol consumption). Each food component is scored from 0 to 10 points and summed to create the overall AHEI score ranging from 0 to 110. Higher AHEI scores are associated with a lower risk of coronary heart disease and diabetes^26^.

### Oral bacterial nitrite generating versus depletion gene abundance

Details of the sample collection, bacterial assessment, taxa identification of the oral microbiome, and estimation of the functional gene profiles from 16S rRNA marker gene sequences have been previously published^11^ and is summarized in the Supplemental Materials. Briefly, subgingival plaque samples (6–8 teeth per participant) were collected from predefined index teeth using sterile curettes and pooled by shallow (probing depth of <4 mm) versus deep (probing depth ≥4 mm) collection sites. Microbial DNA extraction and PCR amplification targeting the V3-V4 region of the 16S rRNA gene (using 341F/806R universal primers) were conducted and underwent next generation sequencing on a MiSeq (Illumina) platform. Following a data curation pipeline in QIIME, the bioinformatics tool PICRUSt2 was used to estimate metagenomic content from the 16S rRNA sequencing, and a weighted average of the within-mouth gene abundance values across pooled samples from both deep and healthy sites used. Our previous analysis found that the ratio of gene abundance values encoding for bacterial reductases along the NO_3_-NO_2_-NO pathway that form nitric oxide (NO) versus ammonium/ammonia (NH_3_) from nitrite (i.e. nitrite reductases that form NO (*nirK & nirS)* in the numerator, and nitrite reductases that form NH_3_ (*nirA, nirB, nirD, nrfA & nrfH*) in the denominator), and thus representing enhanced net capacity for nitrite and nitric oxide generation, was most consistently associated with lower cardiometabolic risk^11^.

Therefore, we focus on this *a priori* determined gene abundance summary ratio which we refer to hereafter as the **M**icrobiome **I**nduced **N**itric oxide **E**nrichment **S**core or MINES.

### Statistical Analysis

Multivariable linear regression analyses regressed composite cardiometabolic risk Z-score and individual cardiometabolic risk biomarkers on tertiles of energy-adjusted total dietary nitrate intake. For all models, we adjusted for total energy intake (kcal) when using the energy-adjusted nitrate intake^22^. Adjusted models (Model 2) controlled for the potential confounders: sex, age, race and ethnicity, education, smoking status, and BMI, and additionally for adherence to the AHEI diet score in the fully adjusted model (Model 3).

Means of the cardiometabolic risk biomarkers are presented in tables across tertiles of dietary nitrate intake. As a more rigorous form of confounder control, sensitivity analyses in healthy participants without hypertension (as per the 2017 American Heart Association criteria^27^: SBP <130 mm Hg, or DBP <80 mm Hg, or self-reported hypertension diagnosis) or prediabetes (using the American Diabetes Association criteria: i) Fasting plasma glucose ≥100 mg/dl and <126 mg/dl; or ii) Haemoglobin A1c (HbA1c) ≥5.7% and <6.5%)^28^, were performed (n=462).

To assess the interaction between the microbial functional profile and dietary nitrate intake, the MINES variable was added to the multivariable models above along with a MINES*dietary nitrate intake interaction term. When assessing interaction, periodontal status was additionally adjusted for as a potential confounder of the oral microbiome to cardiometabolic risk association in multivariable models. Means of cardiometabolic risk biomarkers across median split of dietary nitrate intake were presented in strata defined as above vs. below median MINES and vice versa. The X^2^ test for the interaction term was assessed as significant at an α=5% level.

## RESULTS

### Participant characteristics and dietary nitrate intake

Demographic characteristics of the cohort are shown in Table 1. The study population had a mean age of 31±9 yrs and 73% were female. Moderate and Severe periodontitis was present in 26% and 2% of participants respectively, and a majority (79%) had at least a bachelor’s degree and were overwhelmingly never smokers (87%). The mean AHEI dietary pattern score of 46±12 was comparable to previous reports for a US population^26^. In our population, median [IQR] and mean [SD] dietary nitrate intake was 143 [72, 245] mg/day and 190±171 mg/day respectively. The major contributors to mean daily dietary nitrate intake include cooked spinach/greens (37%), lettuce (19%), and raw spinach/greens (9%), the remainder arising from other plant foods (Table S2). The top contributing items to dietary nitrate intake did not differ by tertile of dietary nitrate intake.

**Table 1.** Characteristics of the study participants by tertile of dietary nitrate intake.

|  | **Total**<br>**(N=668)**<br>**143**<br>**[72,245]\***<br>**mg/day** | **Tertile 1**<br>**(n=222)**<br>**55**<br>**[36,72]**<br>**mg/day** | **Tertile 2**<br>**(n=223)**<br>**143**<br>**[120,170]**<br>**mg/day** | **Tertile 3**<br>**(n=223)**<br>**310**<br>**[245, 436]**<br>**mg/day** | **Pvalue\*\*** |
| --- | --- | --- | --- | --- | --- |
| **Age (years)** | 31.3 ±9.3 | 32.3 ±9.5 | 31.1±9.0 | 30.5 ±9.1 | 0.09 |
| **Sex** |  |  |  |  |  |
| Male | 27% | 32% | 28% | 22% | 0.03 |
| Female | 73% | 68% | 72% | 78% |  |
| **Race/ethnicity** |  |  |  |  | <0.0001 |
| Hispanic | 28% | 36% | 26% | 22% |  |
| Non-Hispanic White | 31% | 23% | 40% | 31% |  |
| Black | 14% | 18% | 10% | 13% |  |
| Other | 27% | 24% | 23% | 34% |  |
| **Education** |  |  |  |  | <0.0001 |
| < Bachelor's Degree | 21% | 28% | 17% | 18% |  |
| Bachelor's Degree | 51% | 54% | 46% | 54% |  |
| > Bachelor's Degree | 28% | 18% | 37% | 27% |  |
| **Smoking Status** |  |  |  |  | 0.18 |
| Never | 87% | 85% | 89% | 86% |  |
| Former | 6% | 5% | 5% | 9% |  |
| Current | 7% | 9% | 6% | 5% |  |
| **Periodontitis** |  |  |  |  | 0.05 |
| None/Mild | 72% | 67% | 77% | 73% |  |
| Moderate/Severe | 28% | 33% | 23% | 27% |  |
| **BMI (kg/m<sup>2</sup>)** | 25.4 ±5.9 | 26.0 ±6.3 | 25.4±5.7 | 24.9± 5.6 | 0.12 |
| **Total energy intake (kcal)** | 1760±967 | 1918 ±1165 | 1790 ±877 | 1574 ±793 | 0.0007 |
| **Dietary nitrate intake (mg/day)** | 190±171 | 54 ±21 | 143 ±31 | 371 ±182 | <0.0001 |
| **Alternate Healthy Eating Index (AHEI)** | 46.4±12.1 | 41.1 ±10.3 | 47.2 ±11.9 | 50.9 ±12.1 | <0.0001 |
| **Cardiometabolic Risk Biomarkers** |  |  |  |  |  |
| CMZ | 0.01±4.2 | 0.75 ±4.1 | -0.06±4.2 | -0.7±3.9 | 0.001 |
| Systolic Blood Pressure, mmHg | 116 ±12 | 117 ±12 | 117±12 | 116±12 | 0.48 |
| Diastolic Blood Pressure, mmHg | 72.0 ±8.8 | 72.7 ±9.1 | 71.7±8.9 | 71.5±8.4 | 0.34 |
| HbA1c, % | 5.25 ±0.3 | 5.30 ±0.3 | 5.24 ±0.3 | 5.21±0.3 | 0.02 |
| Fasting Plasma Glucose, mg/dL | 84.4 ±7.5 | 85.8 ±8.0 | 84.2±7.2 | 83.1±7.2 | 0.001 |
| Insulin, $\mu\text{U/mL}$ | 6.2<br>[4.4, 9.3] | 6.9<br>[4.9, 9.8] | 6.0<br>[4.4, 9.7] | 5.8<br>[4.3, 8.8] | 0.04 |
| HOMA-IR<sup>1</sup> | 1.3<br>[0.9, 2.0] | 1.4<br>[1.0, 2.1] | 1.2<br>[0.9, 2.0] | 1.2<br>[0.8, 1.8] | 0.01 |
| **MINES bacterial**<br>**gene abundance**<br>**ratio<sup>2</sup>** | 0.57 ± 0.30 | 0.52 ± 0.28 | 0.60 ± 0.32 | 0.59 ± 0.29 | 0.004 |

Table values are mean ± SD or median [IQR] for continuous variables, and percentages % for categorical variables.
\* Median [IQR] dietary nitrate intake by tertile of dietary nitrate intake.
\*\**P* values are for ANOVA F statistics or $\chi^2$ tests for any differences in level of covariates between participants across the tertiles of dietary nitrate
Missing values: n=13 for smoking status; n=7 for BMI
<sup>1</sup>Insulin resistance as measured by the Homeostasis Model Assessment for Insulin Resistance (HOMA-IR); BMI indicates body mass index; HbA1c indicates haemoglobin A1c.
<sup>2</sup> MINES bacterial gene abundance ratio: Microbiome Induced Nitric oxide Enrichment Score was calculated as a ratio of microbial gene abundances necessary for NO (nitric oxide) vs. $\text{NH}_3$ (ammonia) production. This score is a surrogate for enhanced net capacity for nitrite and nitric oxide generation

Compared to the lowest tertile of dietary nitrate intake, those in the highest tertile were more likely to be female, have better education, and be of “White” or “Other” ethnicity (Table 1). Participants in the two highest tertiles of dietary nitrate intake had a significantly higher MINES summary ratio and higher AHEI score.

### Association of dietary nitrate intake with cardiometabolic risk

Dietary nitrate intake in the third tertile vs. first tertile was significantly associated with lower plasma glucose, plasma insulin, and HOMA-IR in multivariable adjusted models (Table 2, Model 2). While adjustment for adherence to AHEI score attenuated the associations between dietary nitrate intake and cardiometabolic risk biomarkers (Table 2, Model 3), this may be an overadjustment as dietary nitrate intake is strongly correlated with a healthy diet including vegetable intake. Sensitivity analyses in normotensive and prediabetes-free participants (Supplementary Table S3), showed similar results to the full sample, and remained significant even when controlling for adherence to AHEI score.

**Table 2.**
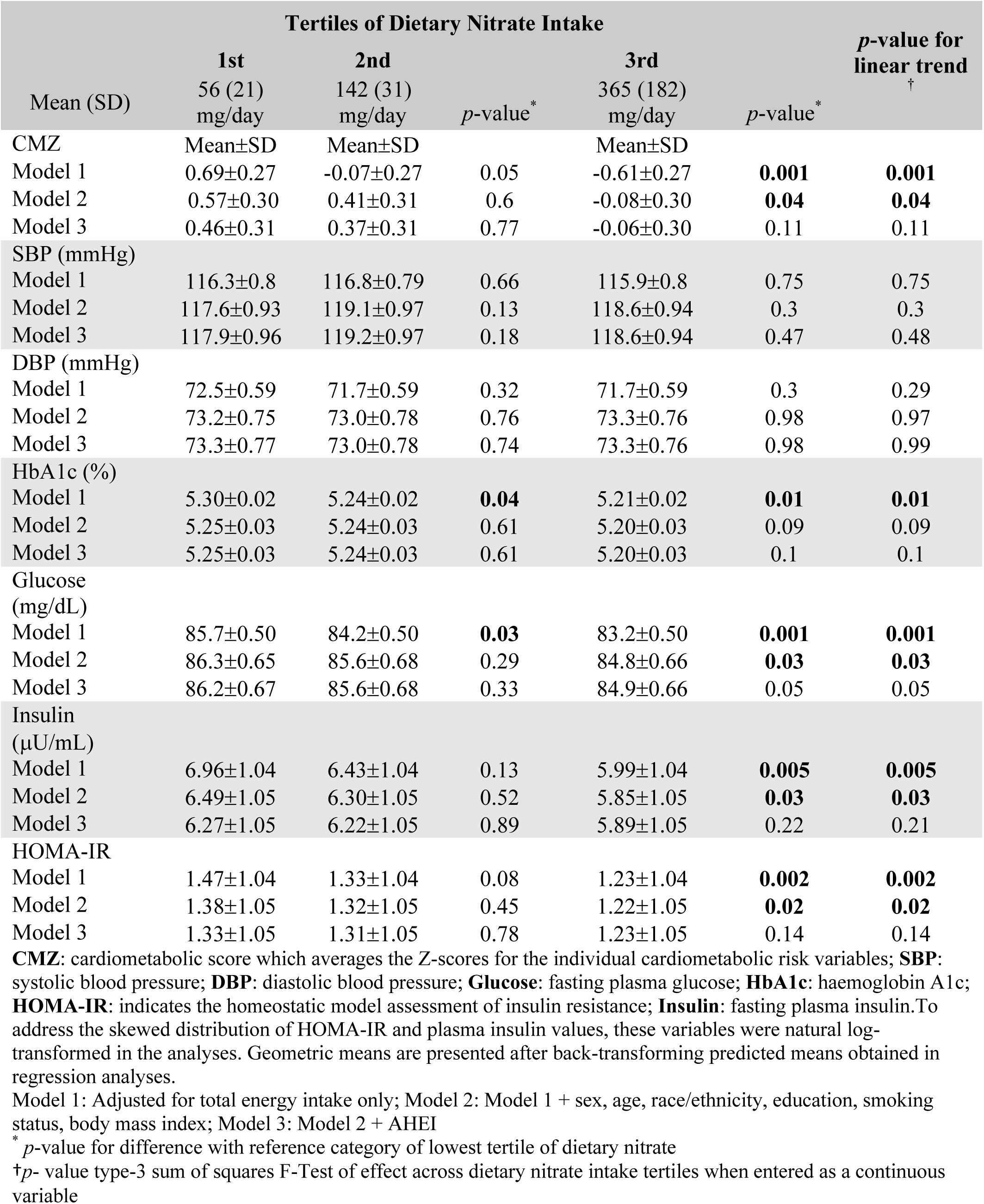
Association of tertiles of dietary nitrate with cardiometabolic risk variables in the full sample (n=668)

### Interaction between microbiome induced nitric oxide enrichment score (MINES) and dietary nitrate intake on cardiometabolic risk

Figure 1B presents the mean cardiometabolic risk biomarker levels across median split of dietary nitrate intake (i.e high (≥median) vs. low (<median)), stratified by high (≥ median) vs. low (<median) MINES. Higher nitrate intake was associated with lower glucose, insulin and HOMA-IR levels among participants with a low MINES in comparison to those with a high MINES. The interactions were statistically significant for insulin and insulin resistance. For example, among participants with a low MINES, mean insulin [95%CI] levels in high vs. low nitrate consumers were 5.8[5.3,6.5] vs. 6.8[6.2,7.5] (p=0.004) while respective insulin levels were 6.0[5.4,6.6] vs. 5.9[5.3,6.5] (p=0.76) among partcipants with high MINES (interaction p=0.02) (Supplementary Table S3). Table 3 presents the same interaction analyses as Figure 1B but from an alternative perspective – when stratified by high vs. lower dietary nitrate intake and with the p-values for high vs. low MINES within dietary nitrate strata. For example, high vs. low MINES was associated with lower mean HOMA-IR [95% CI] levels, 1.2 [1.1,1.4] vs. 1.5[1.3,1.6] (p=0.004) among participants with low dietary nitrate intake, but not in those with high dietary nitrate intake 1.3[1.1,1.4] vs. 1.2[1.1, 1.4](p=0.62)(interaction p=0.02).

**Table 3.** Estimated Mean Cardiometabolic Risk Biomarker (95% CI) Values among Low vs. High MINES, Stratified by Low vs. High Dietary Nitrate Intake.

|  | Low Nitrate Intake* |  | *p*<sup>‡</sup> | High Nitrate Intake* |  | *p*<sup>‡</sup> | P-value for interaction |
| --- | --- | --- | --- | --- | --- | --- | --- |
|  | Low MINES**<br>(n=178) | High MINES**<br>(n=156) |  | Low MINES**<br>(n=156) | High MINES**<br>(n=178) |  |  |
| CMZ | 0.99<br>(0.33, 1.63) | -0.12<br>(-0.79, 0.56) | **0.002** | 0.13<br>(-0.54, 0.80) | -0.07<br>(-0.71, 0.57) | 0.56 | 0.07 |
| Systolic Blood Pressure (mmHg) | 119<br>(118, 121) | 117<br>(115, 119) | **0.02** | 119<br>(117, 121) | 118<br>(116, 120) | 0.19 | 0.49 |
| Diastolic Blood Pressure (mmHg) | 74<br>(72, 75) | 72<br>(70, 74) | 0.07 | 74<br>(72, 76) | 73<br>(71, 74) | 0.19 | 0.73 |
| HbA1c (%) | 5.25<br>(5.19, 5.31) | 5.23<br>(5.17, 5.29) | 0.60 | 5.23<br>(5.16, 5.29) | 5.20<br>(5.14, 5.26) | 0.39 | 0.81 |
| Glucose (mg/dL) | 87<br>(85, 88) | 86<br>(84, 87) | 0.23 | 85<br>(83, 86) | 85<br>(84, 87) | 0.69 | 0.26 |
| Insulin (μU/mL) | 6.8<br>(6.2, 7.5) | 5.9<br>(5.3, 6.5) | **0.005** | 5.8<br>(5.3, 6.5) | 6.0<br>(5.4, 6.6) | 0.64 | **0.02** |
| HOMA-IR | 1.5<br>(1.3, 1.6) | 1.2<br>(1.1, 1.4) | **0.004** | 1.2<br>(1.1, 1.4) | 1.3<br>(1.1, 1.4) | 0.62 | **0.02** |

Multivariate linear regression models adjusted for total energy intake, sex, age, race/ethnicity, education, smoking status, body mass index, periodontal status and Alternative Healthy Eating Index
\*Median nitrate intake [25<sup>th</sup>, 75<sup>th</sup> percentile] = 143 [72, 245] mg/day.
\*\***MINES**: Microbiome Induced Nitric oxide Enrichment Score was calculated as a ratio of microbial gene abundances necessary for NO vs NH<sub>3</sub> production. This score is a surrogate for enhanced net capacity for nitrite and nitric oxide generation. Low vs. High MINES was defined by overall median split of MINES in the full population. Median MINES [25<sup>th</sup>, 75<sup>th</sup> percentile]= 0.54 [0.37, 0.74].
†P-value for difference between groups with above vs. below median MINES within strata of dietary nitrate intake.

## DISCUSSION

In this cross-sectional analysis of young, diabetes-free participants, we found higher dietary nitrate intake to be associated with a lower cardiometabolic risk score which was driven by lower levels of fasting glucose, insulin, and insulin resistance, especially among normotensive and prediabetes-free individuals. However, more importantly, through interaction analyses we observed that higher nitrate intake was associated with lower insulin and HOMA-IR but only among participants with low abundance of oral nitrite enriching bacteria. Similarly, enrichment for oral microbiome-dependent NO generation was most strongly associated with lower cardiometabolic risk among individuals with low dietary nitrate intake, suggesting that the oral microbiome could buffer against the potentially deleterious impacts of low nitrate consumption as illustrated in Figure 1A.

This study advances the existing literature investigating the association of dietary nitrate intake with cardiometabolic health outcomes^29^, by extending beyond blood pressure outcomes to include metabolic risk markers related to impaired glucose regulation. The findings of higher dietary nitrate intake associated with higher levels of early metabolic risk biomarkers such as insulin resistance, and plasma insulin, are supported by experimental animal trials demonstrating similar beneficial metabolic effects^8,30^. However, human trials and studies have been less conclusive^30^, with several studies observing a lack of effect of dietary nitrate on glucose hemostasis, insulin signaling^35^, or incident diabetes^4,5^. Key differences in population characteristics of prior studies, for example as reported by Bahodoran et al. 2017^5^, such as a substantially higher dietary nitrate intake (mean ± SD: 434 ±147 mg/day vs. 190 ±171 mg/day) and rates of smoking (22% vs. 7%) compared to our study population, may explain the different associations previously observed. Our dietary nitrate intake was 143 [72, 245] mg/day, modestly higher than the nitrate intake (median=108 mg) across 51 studies from different countries^31^, but comparable to the mean dietary nitrate intake (152 mg/day) observed in a previous US-based cohort^32^, and in a similarly young French cohort (198 mg/day)^4^. Other studies have suggested that the beneficial effects of increased dietary nitrate intake may plateau beyond ∼60 mg of dietary nitrate intake (∼1 cup of leafy vegetables)^33,34^, therefore, it is possible that nitrate intake exceeding biological thresholds does not add extra cardiometabolic benefit. Alternatively, the influence of smoking on systemic nitrite levels could have blunted the beneficial effects of nitrate intake on metabolic biomarkers in previous studies^29,35^. Differing population characteristics could have also partially explained our somewhat surprising observation that dietary nitrate levels were not related to improved blood pressure, which is inconsistent with some prior studies^3,33,34^.

Our study population is distinctly younger than previous study populations, with a mean age of only 31 years compared to other studies with a mean age of greater than 50 years old^29^.

Some trials have reported greater effects of nitrate supplementation on blood pressure in older adults^36,37^ who might have less capacity for NO-generation and blood pressure control through the canonical L-arginine-nitric oxide pathway, and this unexpected finding in our cohort could reflect the modulating effect of age on the association between dietary nitrate intake and cardiometabolic outcomes.

Crucially, our interaction analysis aligns with the idea that the benefits of nitrate consumption are partially dependent on the oral microbiome, and this omission in prior studies could also partially explain inconsistent prior findings in nitrate supplementation trials and observational studies. The observation of higher nitrate intake associated with better cardiometabolic outcomes only in those with low MINES could also explain the previously observed plateauing of effects of increasing nitrate intakes^33,34^. Large variations in the net nitrate and nitrite-generating capacity of the oral microbiome between individuals have been observed^12,38^. Several oral bacteria along the NO_3_- NO_2_- NO pathway, primarily in the genera *Neisseria*, *Haemophilus*, *Granulicatella*, *Veillonella*, *Prevotella*, *Corynebacterium*, *Actinomyces*, and *Rothia*^39,40^, have been identified as associated with cardiometabolic health, and higher levels of oral nitrate-reducing capacity and net nitrite generation are associated with lower cardiometabolic risk^2,11,12^. This corresponds with the results of a recent large (n=8973) cross-sectional investigation, which found a link between gut microbiota and coronary atherosclerosis; interestingly, several of the taxa observed to relate to atherosclerosis were oral species commonly originating from the mouth. Moreover, bacterial metabolic functional pathways related to dissimilatory nitrate reduction were implicated^41^.

Our current findings provide more explicit support for the significance of the bacterial nitrate metabolism pathway and its association with cardiometabolic health, by studying the oral microbiome and its interaction with nitrate intake directly.

Prior research has shown that interventions which adversely modulate the oral microbiome, such as mouthwash use, can blunt the effects of experimental nitrate supplementation on circulating nitrite levels, blood pressure^9,10^, and markers of glucose regulation^9,42^.

Additionally, while smoking is known to affect plasma concentrations of nitrate and nitrite^35^, smoking has also been associated with alterations to the oral microbiome^43^. Smoking-induced alterations of the oral microbiome is characterized by a depletion in the relative abundance of Proteobacteria and an enrichment of Firmicutes. Prior work in the ORIGINS cohort has found the relative abundance of Proteobacteria to be related to lower levels of inflammation and cardiometabolic biomarkers while the reverse was true for abundance by Firmicutes^44^. If smoking also influences the abundance of taxa with net nitrite-generating capacity, it is possible that the influence of smoking on cardiometabolic risk might be partially mediated via the effect on the nitrite-generating microbiome; a topic for which future research is required.

Both dietary nitrate supplementation and nitrate-reducing probiotics have been suggested as possible interventions leveraging the enterosalivary pathway of NO generation^45^, but when to use either is less clear clinically. Firstly, our findings inform the amount of nitrate consumption potentially relevant to cardiometabolic health. The median split of low and high nitrate intake used in the interaction analyses of 143 mg, and the difference in median nitrate intake in tertile 3 (310 mg/day) vs. tertile 1 (55mg/day) of ∼250 mg/day, is on the low side of nitrate doses used in intervention trials which ranged from 155 mg to as high as 1100 mg^1^.

This suggests that even modest increases in nitrate consumption reflective of real-world intakes (as a component of vegetables consumed) might have cardiometabolic benefits. In fact, prior observational studies reported that increases in dietary nitrate intake as low 30 mg/day may have cardiometabolic benefits^34^. Secondly, as suggested by our interaction analyses, an increase in dietary nitrate intake may be most beneficial for participants with low MINES. Previous experimental studies have demonstrated that dietary nitrate can also act as a prebiotic^46^, encouraging the growth of oral nitrite-generating bacteria^37,47^ within hours following nitrate supplementation^39^. Correspondingly, in this study, we found that dietary nitrate intake was associated with higher net nitrite-generating capacity of the oral microbiome MINES at a population level as well, suggesting that habitual high nitrate intake environments may provide selective pressures for oral bacterial MINES. What proportion of cardiometabolic benefit observed through increased dietary nitrate intake is caused by an increase in MINES versus increased salivary nitrate to be reduced, or both, remains to be tested in future well-designed prospective clinical trials. Finally, compliance to a change in dietary patterns is a common challenge facing the translation of proven dietary interventions into population-based settings. Notably, our finding that elevated MINES values were more strongly related to beneficial cardiometabolic outcomes among those with low nitrate consumption, suggests that probiotic oriented interventions could be most beneficial in populations more resistant to dietary behavior changes. This finding is supported by a recent study showing significantly increased nitrite production, comparable to that produced by a 10-fold higher nitrate supplementation, when adding a *Rothia aeria* probiotic to low levels of nitrate supplementation^45,48^.

### Study Limitations & Strengths

This study has some limitations. We utilized cross-sectional data, and temporality of the variables cannot be ascertained. The assessment of diet and oral microbiome was measured at only at a single time, and is subject to change over time and the lifecourse. While we controlled for several demographic and lifestyle factors, the possibility for residual confounding remains. We are unable to fully separate the potential effects of dietary nitrate intake from the many other nutrients (e.g. Vitamin A, C, E, folate, dietary fiber) that are beneficial for cardiometabolic outcomes and are also found in a high vegetable diet^34^.

Similarly we could not account for the interplay of nutrients such as Vitamin C and thiocynate produced from cruciferous vegetables, which has been shown to interact with dietary nitrate when consumed together, blocking or promoting NO generation^29^. Although we utilized a validated US database for nitrate content, and supplemented this with newer databases to comprehensively calculate total nitrate intake, measurement error is still likely as the nitrate content of vegetables can vary substantially depending on growth factors and cooking preparation methods^13^; nitrate intake from drinking water also was not assessed.

Nevertheless, as our study population mainly resides within the NYC Tri-state area, these possible measurement errors in dietary nitrate intake are likely to be non-differential to the outcome of interest, biasing the effect towards the null.

Limitations arising from our bacterial measures have been detailed elsewhere^11^. Although the tongue microbiome is thought to be the primary site for oral nitrate reduction, our current study utilized subgingival plaque samples for analysis. Nevertheless, while these sites are compositionally distinct, the genera often identified as relevant to the enterosalivary NO generation and shown to change with nitrate supplementation (e.g. increased *Rothia, Nesseria* and decreased *Prevotella)*, exhibit substantial overlap between subgingival plaque^49^, saliva^46^, or tongue^38^ microbial communities. We have not accounted for local environmental factors in the oral cavity, including oral pH, oral diseases aside from periodontitis, or mouthwash use.

Due to the cross-sectional nature of this study, it is also possible that the presence of high MINES in those with a low dietary nitrate intake is indicative of robust endogenous NO formation through mammalian NO synthase and the canonical L-arginine-nitric oxide pathway. The high NO formed is quickly oxidised to NO_3_ and recycled as high salivary nitrate concentration which promotes the growth of high levels of MINES, and increased salivary nitrate reduction and salivary nitrite formation. Thus endogenous NO production could be confounding the association observed, and our findings could reinforce that dietary nitrate supplementation is most effective in those with impaired or lower endogenous NO production, rather than that directly indicating that increasing MINES levels is beneficial.

Therefore, future studies should assess the direct effect of a probiotic to increase MINES on salivary nitrite formation, while controlling for the baseline levels of endogenous NO production and dietary nitrate intake. Nevertheless, MINES may still be useful as a predictive biomarker as endogenous NO production capacity is difficult to measure directly. Finally, the functional capacity of the oral microbiome was inferred from 16S rRNA gene sequences, and future studies using metagenomic shotgun sequencing are needed to validate the utility of microbial gene abundances in predicting cardiometabolic risk.

Despite these limitations, this study has notable strengths. ORIGINS consists of a diverse study population and measured a robust set of risk factors to minimize confounding. We utilized a comprehensive FFQ, and a validated US-specific nitrate database most relevant to our population, supplemented by more recent dietary nitrate databases to capture a broad spectrum of dietary nitrate intake. The young, diabetes-free study population limits reverse causation, and enabled us to examine the interplay between dietary nitrate intake, the oral microbiome, and cardiometabolic biomarkers before the development of clinical disease and related pharmacological interventions and behavioral changes.

## CONCLUSIONS

We found higher dietary nitrate intake was only associated with lower glucose, insulin and insulin resistance among individuals with reduced capacity for oral microbe-induced nitrite enrichment, in a young, generally healthy population. If these findings represent a causal diet*microbiome interaction on cardiometabolic health, they have important implications for precision nutrition oriented research as outlined in the *2020-2030 Strategic Plan for NIH Nutrition Research*^14^. These findings also have implications for the development of beneficial pre/probiotics as our results suggest that increasing the nitrite-generating potential of the oral microbiome might buffer against poor cardiometabolic health among individuals with low nitrate intake. Larger, longitudinal studies, with more precise measures of dietary nitrate intake and measures of the tongue metagenomic capacity, will provide a deeper understanding of the potential interaction between the oral microbiome and dietary nitrate intake in relation to cardiometabolic outcomes.

## Data Availability

Available upon request to the corresponding author

## Acknowledgements

We thank the participants and study staff of ORIGINS for their valuable contributions.

## Sources of Funding

This research was supported by NIH grants R00 DE018739, R21 DE022422 and R01 DK 102932 (to Dr. Demmer). Dr. Demmer also received funding from a Pilot & Feasibility Award from the Diabetes and Endocrinology Research Center, College of Physicians and Surgeons (DK-63608). This publication was also supported by the National Center for Advancing Translational Sciences, National Institutes of Health, through Grant Number UL1TR001873. Rebecca Molinsky was supported by institutional training grant T32HL007779 from the National Institutes of Health.The content is solely the responsibility of the authors and does not necessarily represent the official views of the NIH.

## Disclosures

The authors declare no conflict of interest.

## Data Availability Statement

All sequencing data are available through Qiita50 under study ID 14375 for subgingival plaque samples. Raw sequence data is also available through EBI accession PRJEB50261 for subgingival plaque samples.

## Notes

### Competing Interest Statement

The authors have declared no competing interest.

### Clinical Trial

N/A

### Author Declarations

The Columbia University and University of Minnesota Institutional Review Boards approved the protocol. All participants provided informed consent.

